# Healthiness of Foods and Beverages in Outdoor Advertisements in Xela, Guatemala: A Commercial Determinants of Health Interpretation

**DOI:** 10.1101/2024.06.28.24309660

**Authors:** Yulia E. Chuvileva, Aiken Chew, Omni Cassidy, Sophía Dávila, Arie Manangan, George Rutherford, Joaquin Barnoya, Brigette Ulin

## Abstract

This study measured the spatial exposure to outdoor advertising of foods and beverages (and their healthiness) in central Xela, a Western Guatemalan city that is home to 181,000 people, 65% of whom are indigenous. We geotagged outdoor advertisements (ads) for foods and beverages in a square mile of Xela, coding for the modality of the advertisement, the types of items marketed, and their healthiness: “most healthy,” “middle healthy,” or “least healthy.” We observed 92 commercial-grade outdoor food and beverage ads across eight modalities, with vehicles (52%, 48/92), branded storefronts (25%, 23/92), and bus stops (13%, 12/92) the most prevalent. While all kinds of food and beverage businesses promoted their products on vehicles, branded storefronts were a modality almost exclusively used by soda companies, while bus stops were disproportionately used by fast food restaurants. We also identified 21 home-based ads, 86% (18/21) of which denoted households selling items they produce or make. Commercial-grade ads promoted largely least healthy foods and beverages, while home-based ads largely exposed people to most and middle healthy items. We interpret the results through the Commercial Determinants of Health Framework.

## Introduction

### Commercial Determinants of Health

Globally, poor diets are responsible for more deaths than any other health risk factor (including smoking), and improvements in unhealthy food and beverage consumption could prevent one in five mortalities around the world [1]. The retail food environments that people rely on to buy foods and drinks is one of five major social determinants of health [2]. Commerce, in turn, drives food and beverage availability, pricing, and marketing.

The commercial determinants of health (CDoH) framework considers both positive and negative impacts of commerce (e.g., business activities, products) on health, based on whether the involved goods and practices are health promoting or health damaging [3]. CDoH scholars also monitor businesses’ political activities, especially in the food and beverage sector [4], to track commercial influences on policy, policy gaps, and the intersections between commercial, political, and social determinants of health [5, 6].

### Outdoor Advertising of Foods and Beverages

Marketing is a key commercial activity within CDoH. Advertisements (ads) drive purchasing and consumption behaviors by priming people with cues to consume specific goods or brands [7]. Different forms and levels of exposure to advertising have been shown to directly affect individuals’ food attitudes and food purchasing and lead to greater consumption of advertised foods and overall calories, particularly by children [8].

Reliable estimates of food and beverage sector marketing expenditures and which groups it reaches in non-higher-income countries are rare. Higher-income country data, thus, provides a useful comparison. In the U.S., for example, food, beverage, and restaurant companies spend U.S. $14 billion per year on advertising (averaging over $42 per person), 80% of which promotes unhealthy items [9]. Food companies have been found to advertise more frequently using specific media in lower-income and middle-income countries than in higher-income ones [10], indicating that the food and beverage industry could be spending similar or higher *per capita* amounts in other countries as in the U.S. Meanwhile, a 2021 review concluded that “children from minority and socio-economically disadvantaged backgrounds are disproportionately exposed to unhealthy food advertising” [11], a result based on available research on the topic, which has, to date, only been done in higher-incomes countries of Australia, England, New Zealand, and the U.S. This evidence suggests that people in different countries and from different groups within countries experience differential levels of exposure to unhealthy food and beverage marketing.

Compared with advertising research on viewing and listening platforms (e.g., film and television, radio, social media), little evidence exists on the healthiness of foods and beverages advertised in outdoor spaces[12]. Yet, outdoor ads are important to study because they spatially expose people to marketing messaging as they go about their typical day. Indeed, studies from Ghana, South Africa, and the U.S. show that outdoor food and beverage ads tend to be placed in high (foot)traffic areas, such as billboards on roadways and posters at bus, metro, and train stations [13-15]. In Australia, Canada, Mexico, New Zealand, the United Kingdom, and the U.S., ads for unhealthy items have also been shown to disproportionately target lower income areas and neighborhoods with a higher proportion of Black, Indigenous, and people of color, as well as outdoor areas in and around places of study frequented by children and youths [11, 16-18].

Public health and applied geographical research has yielded important insights about the distribution and concentration of outdoor ads across some modalities and some population sub-groups in wealthy contexts. Of the 53 studies that exist, most focus on higher-income countries, a single commercial-grade advertising modality (such as billboards or bus stop posters), a single type of food or beverage (like soda), or a specific locational target, such as food and beverage ad concentration around schools [8]. However, no studies have measured exposure to all outdoor food and beverage advertising across multiple modalities in a single geographical area, and few have looked at any modalities in less wealthy countries.

The paucity of research on outdoor food and beverage advertising as a CDoH may contribute to the lack of effective policies on outdoor food marketing[19]. This leaves an actionable policy gap since food and beverage industry self-regulation is often ineffective [20]. Meanwhile, a substantial portion of outdoor food and beverage advertising happens in public areas that governments can more easily regulate [21].

### Case study: diet and health in Guatemala

Over the last several decades, industrially produced and processed foods and beverages have increasingly, but incompletely and unevenly throughout the population, replaced subsistence and freshly prepared items in Guatemala [22]. The marketing and sale of food and beverage brands have facilitated this nutrition transition, which is especially evident in the large and growing *per capita* consumption of sodas and other sugar-sweetened beverages (SSBs) [23]. The nutrition transition has also been facilitated by increased imports of unhealthy items following the integration of Guatemala into regional and global markets through several rounds of trade liberalization after the 1996 signing of the Peace Accords that officially ended the 36-year Civil War [24].

Yet, the nutrition transition in Guatemala has not seen the accompanying epidemiological transition seen in wealthier countries: from infectious diseases and malnutrition to chronic diseases. Instead, the country has seen a compounding of infectious and chronic health problems that disproportionately affect indigenous people. Indigenous children have a stunting prevalence of 60%, compared with 36.4% among non-indigenous groups [25]. And while in 2016 the national diabetes prevalence reached 7.5% [26], research around the same time in Mayan towns of Lake Atitlán found 13.6% diabetes prevalence (and an additional 13.6% pre-diabetes prevalence) [27]. Overall, poor diets are a risk factor for seven of the top ten causes of death in Guatemala [28]: heart disease, diabetes, kidney disease, cirrhosis, stroke, stomach cancer, and diarrheal diseases.

### Employment, educational, and informational context of Guatemala

While Guatemala is an upper-middle-income country, is Central America’s largest economy, and is one of the strongest economies in Latin America, its resources are vastly unevenly distributed. Contrasted to the 52% of the population who lives in poverty [29], the combined wealth of the country’s 260 millionaires amounts to 56% of the country’s GDP [30]. A substantial portion of the individual and corporate wealth has been amassed through the production, distribution, marketing, and sale of industrial foods and beverages.

Outdoor food and beverage advertisements likely play a large CDoH role in Guatemala due to the country’s employment, linguistic, and technological context. At the national level, 65% of the population’s work is unregulated, including farming and small-scale indoor and outdoor retail; a third of the population is employed (largely outdoors) in agriculture; and a fifth of the population does not read or write [31-33]. When it comes to communication, Guatemalans speak 24 different languages, with multiple sub-variations of Mayan languages. While 79% use cell phones, only 45% use computers, and 54% use the internet, with only half the population on social media [34]. As a result, physical forms of advertising on billboards, posters, and signs, especially outdoors, likely remain one of the more reliable ways that advertisers can reach many Guatemalan consumers (with television and radio among the other important modalities).

Outdoor advertising may be especially important for reaching indigenous and peasant communities within Guatemala who are over-represented in outdoor, unregulated work [31], but under-represented in internet and digital media usage [35]. These inequities stem partly from ongoing legacies of historic racism, discrimination, and marginalization of Guatemala’s indigenous and peasant people, including a brutal 1980s genocide against indigenous civilians [36].

This context makes the indigenous-majority city of Xela [shey-lah], Guatemala, an important case study. [Xela is the widely used shorthand for Xelajú [shey-lah-who], the pre-conquest Maya K’iche’ name for the city administratively named Quetzaltenango. We follow others [37] in using the term Xela (instead of Quetzaltenango) to center Guatemalan people’s lived experiences in our writing]. Xela is an important case study because it is an exemplar of many parts of the world where outdoor advertising on physical billboards, posters, and signs remain important commercial strategies. That is, contexts where large proportions of the population: 1) rarely access social media due to cost, literacy, or language barriers; 2) travel by public transport; and 1. 3) work, especially informally, outdoors in agriculture or in ambulatory retail. Xela is thus worthy of study as an important commercial center in Western Guatemala that is home to 181,000 people, 65% of whom are Mayan [38]. The city also draws workers and visitors from the more than 600,000 residents of the department of Quetzaltenango, where 83.5% of the population is informally employed and where 95% of commercial businesses focus their activities on urban areas, especially the city of Xela [39].

Since the CDoH framework and available research evidence shows outdoor food and beverage advertising to be a commercial activity that shapes environments and influences health behaviors and outcomes, our study of Xela, Guatemala aimed to answer the following research questions:

1. What is the total exposure to outdoor food and beverage advertisements in central Xela?
2. What modalities of outdoor food and beverage advertisements exist in Xela?
3. Which entities, businesses, and brands advertise through outdoor modalities in Xela?
4. What is the healthiness of the items advertised outdoors in Xela?

## Materials and Methods

The GIS mapping reported in this paper was one part of a larger, year-long, embedded, ethnographic study that also entailed participant observation and over 120 in-depth interviews with Xela’s consumers, farmers, food retailers, and non-profit and government representatives working to improve the city’s food system. The present paper builds on our accompanying publication detailing Xela’s retail food environment [40].

In late summer 2017, starting with Xela’s Central Park, which anchors a busy residential, commercial, and leisure center of the city, adjacent streets were surveyed on foot in an area of 1.702 km^2^. GPS Kit (Garafa, LLC, Provo, UT) was used to geotag all forms of food and beverage retail and advertising visible from the streets during the mapping periods over several days. Every business, home, street-based advertising structure, and passing vehicle were examined for food and beverage retail or advertising content. This excluded *in-store* point of sale promotions, such as small posters or branded stickers on the insides of store doors or on store equipment, that were visible when passing them by. Such in-store promotions were ubiquitous across the 377 brick-and-mortar retailers and across a portion of the 175 outdoor vendors who had a semi-permanent stall structure that our larger, geographical retail food environment study reported in Xela [40]. The discussion outlines the implications of this methodological decision. While vehicle-based promotions were added as data points, vehicle identifiers were not collected. While the largest and busiest thoroughfares were all captured in the dataset, along with many of the smaller roads, time and resource constraints prevented every single one of the smallest residential streets and cul-de-sacs from being mapped.

Foods and drinks advertised outdoors were coded by: a) ad modality, using some categories found in existing literature (e.g., billboard, bus stop, poster) and adding new ones that had not been reported before (i.e., vehicle-based, large signs, hanging signs, and in-person promotions; and b) the food or beverage items being advertised and/or the brand. All data points were additionally coded for two types of advertisements that emerged from the data: 1) commercial-based ads, defined as professionally designed and printed logos, images, and messaging on paid-for outdoor media, like billboards and bus stops; or 2) home-based ads of hand-written or printed signs hung in residential windows or on the outsides of doors to advertise items for sale in homes.

To assess the healthiness of the food and beverage advertisements in a culturally responsive way, we followed the expert rater methodology detailed in our accompanying manuscript [40]. Four raters determined the advertised items’ healthiness drawing on our database of 174 food and drink items commonly available for sale in Xela. The 174-item list was generated by analyzing data we collected on the consumable items available for sale from central Xela’s indoor and outdoor food retailers we surveyed as part of the larger study of the city’s food environment [40]. While the list is not exhaustive, it represents the most common food categories available for sale in the city, such as egg, bean, and tortilla meals, soups, burger meals, pastas, pupusas, ice-creams, juices, chips, etc. The raters are experts on Guatemala’s food systems and health, including a U.S.-based food anthropologist with a 12-year track record of working in Guatemala and three in-country Guatemalan researchers: an anthropologist of food, a nutritionist, and a physician.

To assess healthiness, the raters first followed prior food retailer healthiness research in Guatemala to code all items either as healthy or unhealthy [41]. However, the raters were unable to complete the task because the foods and drinks sold in Xela did not fit a healthy/unhealthy binary. They then attempted to code the data with a more expansive, three-point relative scale of least, middle, or most healthy, but found it difficult to apply to all 174 items in a single list since many of the foods and drinks are functionally inequivalent. For example, it was difficult to compare the healthiness of raw chicken destined for meal preparation with a freshly-prepared horchata, which are items that people consume under different circumstances and rarely attempt to choose between. To overcome the challenges, the items were separated into two functional categories: 1) “ready-to-consume” (pertaining to the grazing environment) with several sub-categories (snacks, desserts, drinks, meals [breakfasts, lunches, and dinners], and meal accompaniments [e.g., tortillas]) and 2) ingredients (pertaining to the grocery environment), which are items people purchase for preparing meals at home [42]. Each functional category included various variations of available foods or beverages sold in Xela, e.g., the category of desserts included four types of frozen treats, branded industrially produced ice-cream, artisanal ice-cream, chocolate-covered frozen fruit (like *chocobananos*), and frozen yogurts. The four raters then independently rated each of the 174 foods and drinks on a three-point scale of “least,” “middle,” or “most” healthy within their functional categories and sub-categories (see Appendix A), finding the approach feasible due to its contextual relevance (i.e., being based on locally-available food options) and basis in real-world dietary behavior (i.e., how people make food decisions). The advantages of this methodological innovation are discussed in detail in our accompanying manuscript [40]. Each food item’s healthiness rating was then decided by full or majority agreement or resolved by discussion. Most food items (85%, 148/174) received agreement either from all four (53%, 93/174) or three of the four (32%, 55/174) raters, validating the measure as a strong fit for the task. Outdoor ads were then rated for healthiness depending on the foods or beverages they promoted.

## Results

This study found central Xela’s total geographical exposure to outdoor food and beverage ads to be 113 in an area of 1.702 km^2^. A high concentration of ads was located on the main northeastern road into the city from the Pan-American Highway (Figure 1). The busy thoroughfare is a main route for much of the public (buses and microbuses) and private transport through the city.

Figure 1: Distribution of Food and Beverage Advertisements in Xela

**Figure 1 Caption:**
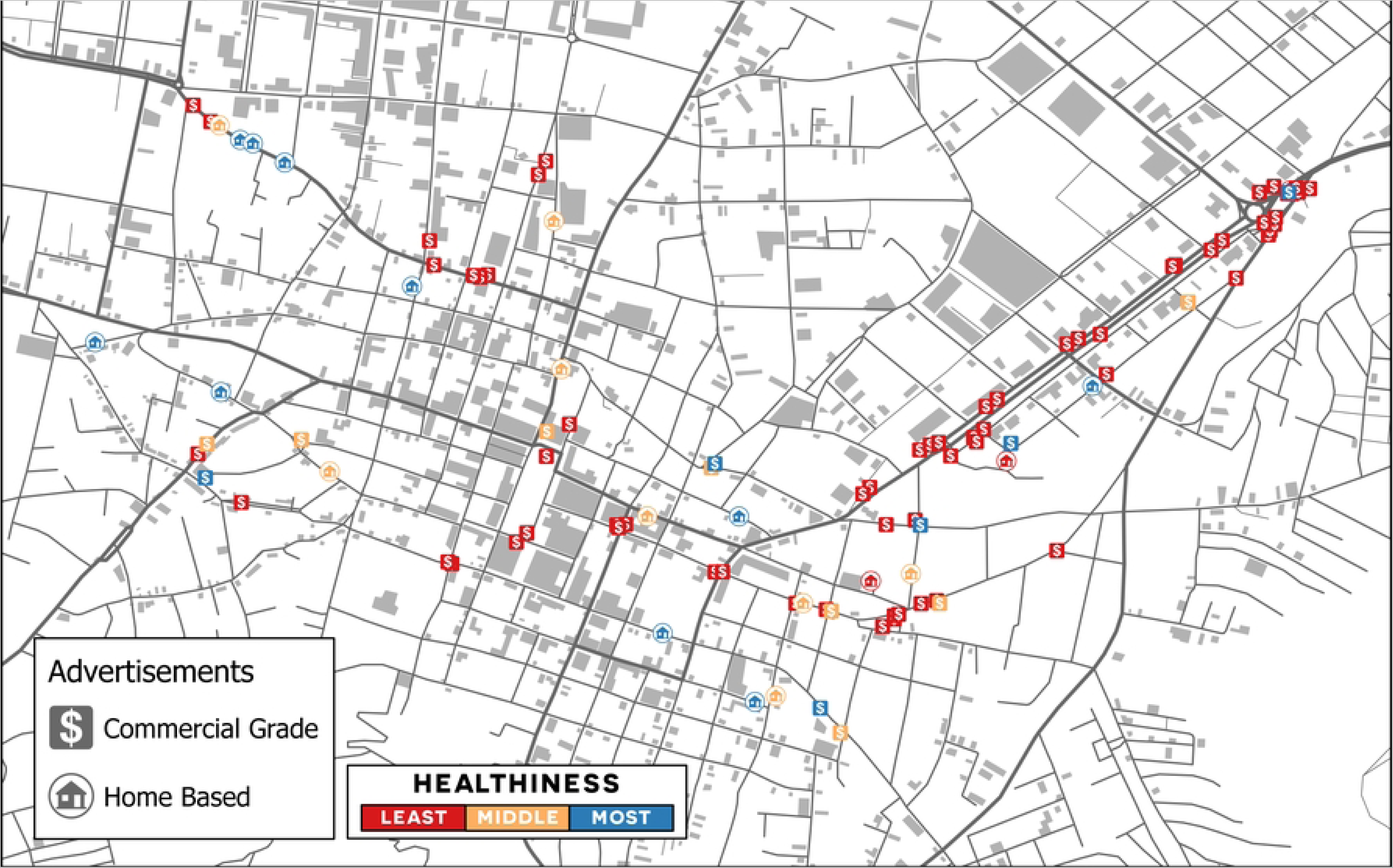
A total of 113 outdoor ads were found in the mapped area of central Xela (1.702 km^2^). A high concentration of ads was located on the main northeastern road into the city from the Pan-American Highway. The busy thoroughfare is a main route for much of the public and private transport through the city.

One modality of home-based ads was found: printed or hand-made signs hung in residential doors or windows. Eight modalities of commercial-grade ads were identified: vehicles (promotional or delivery), branded storefronts, bus stop posters, billboards, large signs (printed canvas banners promoting a product or brand), hanging signs (branded smaller signs), in-person promotions (people dressed in branded clothing promoting brands with flyers and/or mascots), and fixtures (referring to a large, branded, plastic three-dimensional pizza box perched on top of a tall supporting pole).

### Commercial-grade ads

Ninety-two commercial-grade outdoor advertisements in central Xela were observed across eight modalities, with vehicles (52%, 48/92), branded storefronts (25%, 23/92), and bus stops (13%, 12/92) being the most prevalent.

All but one of the vehicles were delivery trucks, vans, or mopeds, with the exception being a promotional vehicle dedicated to visual and audio announcements to advertise products. Almost a third (29%, 14/48) of the vehicles promoted drinks, namely Coca-Cola and Pepsi soda (17%, 8/48), the leading national beer brand, Gallo beer (8%, 4/48), and Salvavidas bottled water (4%, 2/48), a Coca-Cola subsidiary. Almost a quarter (23%, 11/48) of the vehicle-based ads promoted ultra-processed food (UPF) snack brands, such as Frito Lay, Cheetos, and Tortrix, the nation’s leading chip brand. Another quarter (23%, 11/48) promoted ingredients like Lala dairy and Chimex and Perry processed meats. Nineteen percent (9/48) marketed desserts, including Sarita and Pali Deli ice-cream brands and Gimesa cookies. The smallest category of vehicle-based ads (6%, 3/48) was for fast-food meals at the nation’s leading fast chicken brand, Pollo Campero, and Western Guatemala’s regional friend chicken takeout, restaurant, and grocery chain, Albamar.

Of the 23 storefronts, 22 (96%) were branded by SSB companies, namely the soda brands Pepsi, Coca-Cola, Super Cola, and Crush; the exception was Gallo beer.

Of the bus stop ads, 67% (8/12) were for fast-food restaurant meals, consisting of two international chains, Taco Bell and Dominos, and two Guatemalan chains, Pollo Campero and Albamar. The other third (4/12) of bus stop ads were for Maggi consommé packets, a Nestlé brand.

Of the large sign, hanging sign, fixture, and billboard ads: 50% (4/8) were for fast-food restaurants (Pizza Hut, Dominos, and Pollo Campero); 25% (2/8) for ice-cream brands (POPS and Sarita); and 25% (2/8) for alcohol brands (Gallo and Venado).

In addition, there was one in-person promotion that included a mascot, uniformed staff members, and women wearing Xela *traje* (Mayan clothing whose design, patterns, and colors vary by community) promoting the country’s leading fried chicken takeout brand, Pinulito, a cousin company of Pollo Campero.

In terms of functional food and beverage categories, across commercial-grade ad delivery modalities, 42% (39/92) of the ads were for drinks (non-alcoholic and alcoholic beverages), 17% (16/92) for meals, 16% (15/92) for ingredients (grocery) items, 12% (11/92) for snacks, and 12% (11/92) for desserts (Figure 2). In terms of healthiness, 83% of all commercial-grade outdoor ads (76/92) exposed people to least healthy items, 11% (10/92) to middle healthy items, and 7% (6/92) to most healthy items. Of the least healthy advertised foods and beverages, 89% (68/76) were for highly processed items offered by global and national companies, of which 54% (37/68) were sugar-sweetened and alcoholic beverages, 24% (16/68) were fast-food restaurant meals, and 22% (15/68) were snacks and desserts. While two-thirds (32/48) of vehicle modality ads were for least healthy items, painted storefronts and bus stops advertised exclusively least healthy foods and beverages.

Figure 2: Distribution of Commercial-Grade Advertisements across Functional Food and Beverage Groups and Healthiness

**Figure 2 Caption:**
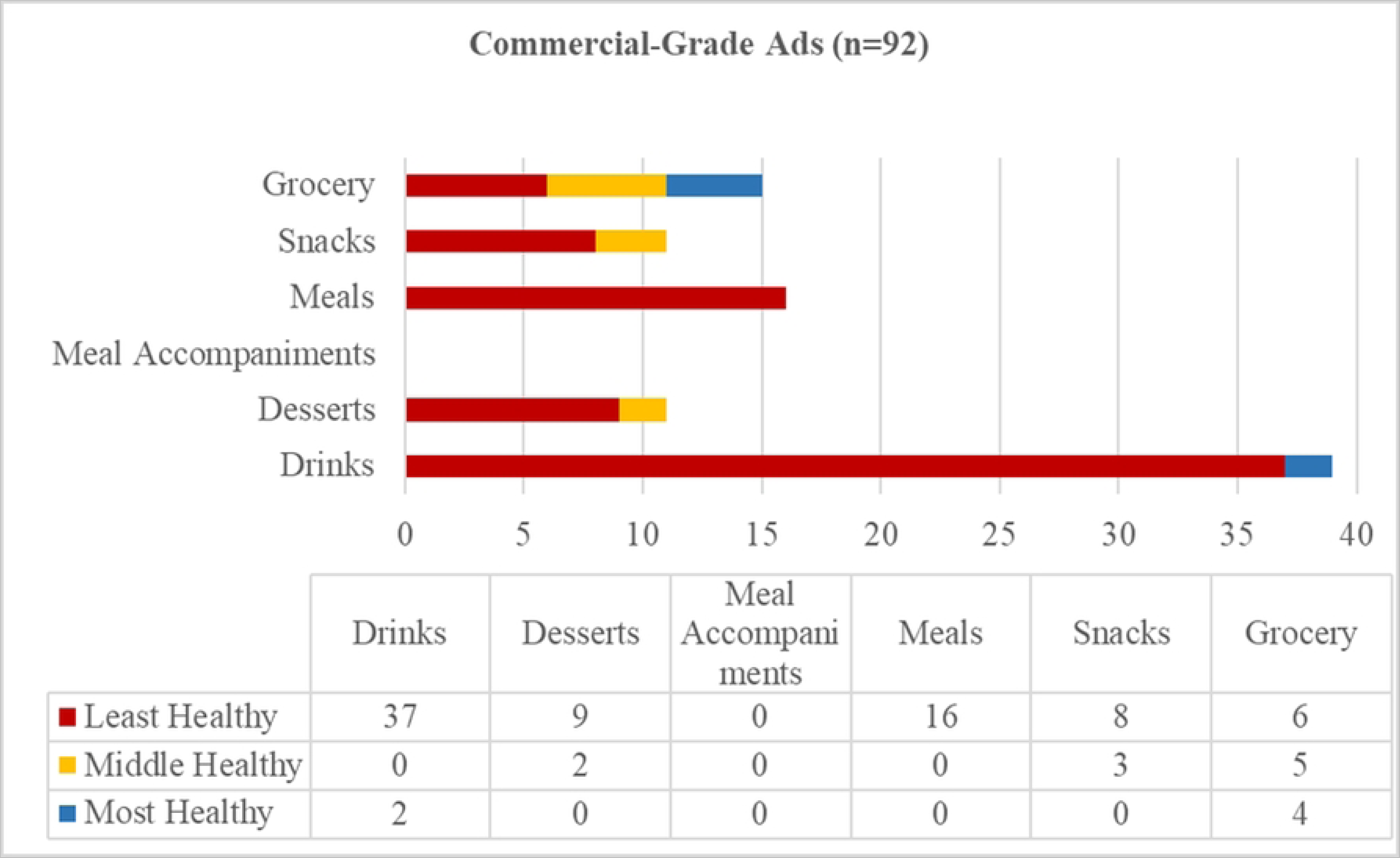
Across commercial-grade ad delivery modalities, 42% (39/92) of the ads were for non-alcoholic and alcoholic beverages, 17% (16/92) for meals, 16% (15/92) for grocery items, 12% (11/92) for snacks, and 12% (11/92) for desserts. In addition, 83% of all commercial-grade outdoor ads (76/92) exposed people to least healthy items, 11% (10/92) to middle healthy items, and 7% (6/92) to most healthy items.

### Home-based ads

Twenty-one home-based ads were observed. Eighty-six percent (18/21) of the signs denoted households selling items they produce or make: ice cream (n=5), chocolate (n=3), *tortillas* (n=2), coffee (n=2), eggs (n=2), *tamales* and *paches* (n=1), honey (n=1), maize (n=1), and *guisquiles* (a native green, pear-shaped squash) (n=1). Fourteen percent (3/21) of the ads were for items being sold on behalf of businesses: alcohol for the national liquor brand, Quetzalteca, (n=1), dairy for the regional business of Lacteos de Patulul (n=1), and donuts on behalf of the brand, American Donuts (n=1). The foods and drinks in home-based ads fell into the following functional categories: desserts 43% (9/21), grocery items 29% (6/21), meal accompaniments 14% (3/21), and drinks 14% (3/21). No snacks or meals were promoted in home-based ads.

Overall, 90% (19/21) of home-based ads promoted the sale of most or middle healthy foods and beverages and 10% (2/21) of least healthy items, with most healthy items appearing in grocery, meal accompaniment, and drinks categories (Figure 3). The smaller sample size of home-based ads (n=21) compared to commercial-grade ads (n=92) prevents any firm conclusions to be made about relative healthiness of items promoted by the two modalities. However, it is noteworthy that the two home-based ads for least healthy foods were those reselling commercial brands of donuts and liquor. Home-based advertisements for foods and drinks that people grew and/or prepared themselves were all in the middle and most healthy categories as determined by expert ratings.

Figure 3: Distribution of Home-Based Advertisements across Functional Food and Beverage Groups and Healthiness

**Figure 3 Caption:**
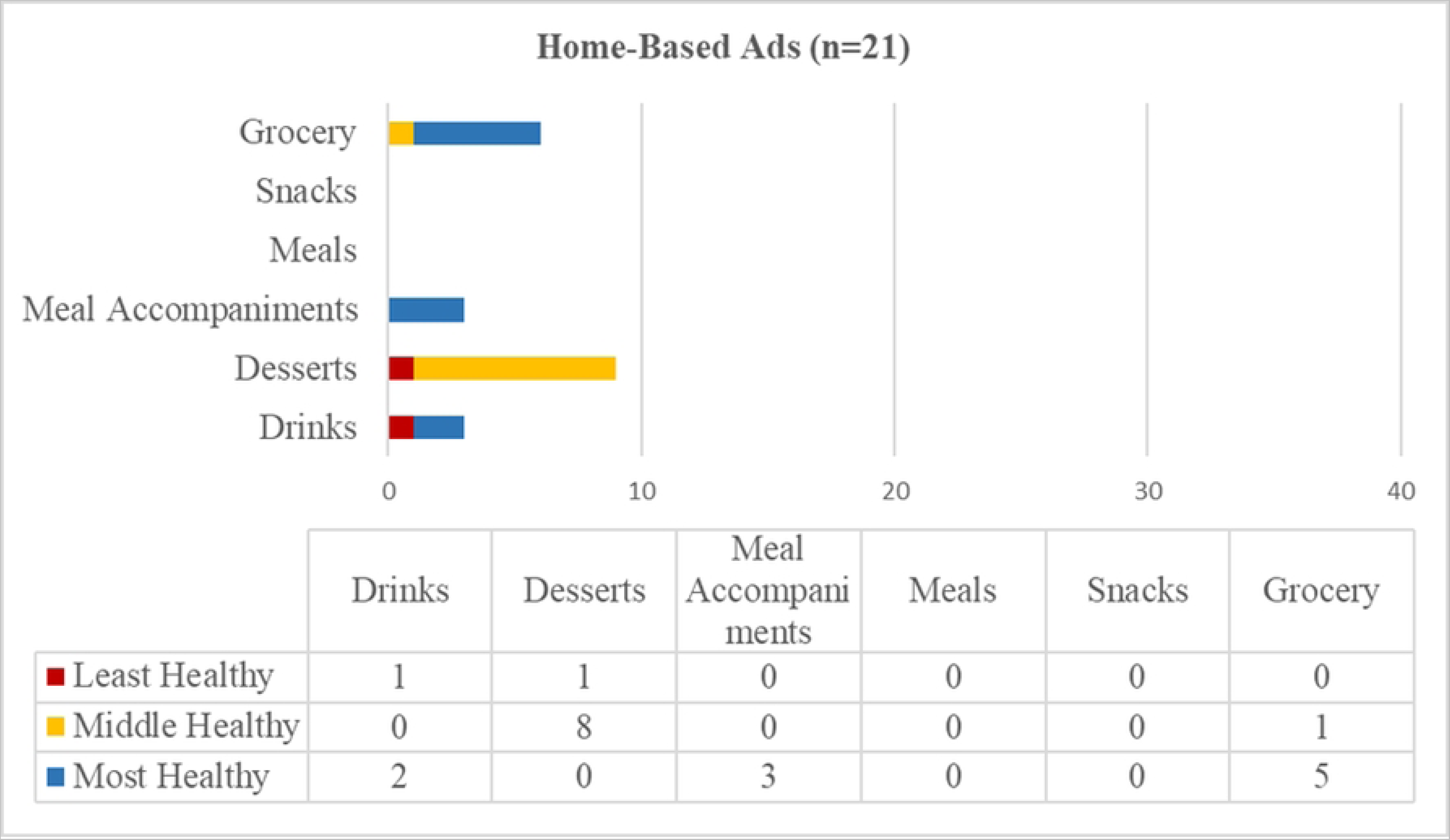
Home-based ads promoted desserts 43% (9/21), grocery items 29% (6/21), meal accompaniments 14% (3/21), and drinks 14% (3/21). No snacks or meals were promoted in home-based ads. Overall, 90% (19/21) of home-based ads promoted the sale of most or middle healthy foods and beverages and 10% (2/21) of least healthy items, with most healthy items appearing in grocery, meal accompaniment, and drinks categories.

## Discussion

This study documented the modalities of outdoor food and beverage advertisements in central Xela; whether people or brands advertise through those modalities; and the healthiness of the items they promote. We identified eight modalities of commercial-grade outdoor ads, including two that are unreported or under-reported in the literature: vehicle-based product promotion and branded storefronts. Together, branded vehicles and storefronts accounted for almost four out of five commercial-grade outdoor food and beverage ads in central Xela. Different businesses dominated different modalities of outdoor ads, with soda companies over-represented in branded storefronts and fast-food restaurants and consommé brands in bus stops, while all kinds of global, national, and regional businesses advertised on vehicles.

In addition, the study identified a second, previously unreported type of advertisements: home-based ads. Home-based ads were dominated not by global and national businesses, but by small-scale, unregulated, non-commercial-grade retail of consumable items, although two of them did resell foods and beverages on behalf of businesses. While making up a fifth of the total observed food and beverage ads, home-based ads tended to be for healthier items than commercial-grade ads. We interpret our findings through the CDoH framework that indicates that retailers engage in both commercial and political activities to increase sales of their products and that CDoH can have both negative and positive impacts on public health nutrition [43].

### Marketing through vehicle delivery

Vehicles were the largest source of exposure to commercial-grade food and beverage advertisements in central Xela, accounting for more ads than all the other commercial-grade modalities combined. Mobile ads were captured from delivery trucks, cars, and mopeds, and via a specialized advertising vehicle. To the best of our knowledge, vehicles as ad modalities have not been previously reported in public health literature that studies food and beverage advertising. This is surprising since such mobile advertising has a long history, with soda companies advertising through branded delivery crates since at least the 1930s [44]. Today, vehicle-based advertising, aka “moving billboards”, is a popular marketing strategy that yields almost 16 million Google results when searching for “publicidad movil Guatemala” (“mobile advertising Guatemala) in Spanish and 83 million results when searching for “branded vehicles for advertising” in English, with many pages in both languages offering vehicle branding services or tips.

The strategy of “making your vehicle fleet work for you” may be attractive to businesses because it is relatively low-cost compared to ads of a similar size, such as billboards. It also may expose many more people to its content since delivery vehicles cover a lot of distance in a targeted way to populations who live close to locations of product retail, and the ads are eye-catching, especially for people walking past or driving through slow or standstill traffic, which is typical of Xela. Mobile ads may constitute an important part of retail food environments that needs further study in terms of levels and effects of exposure, as well as possible public health policy and practice solutions aiming to prevent, reduce, or eliminate any negative impacts.

### Branded storefronts

An additional striking feature of the visual landscape of Xela is entire storefronts and other kinds of buildings painted in brand logos and colors. In some cases, we have observed these branded storefronts inundate the visual sphere of portions of neighborhoods. In the Xela sample, all but one of the painted storefronts advertised soda brands. In this study, painted storefronts (mainly in soda brand colors and logos) were the second largest delivery mode for commercial-grade food and beverage advertising, accounting for more than a quarter of all observed ads in the category. Yet, this advertising modality has been scantly reported in the literature. SSB-company sponsored kiosks have been noted in Guatemala’s schools [45] and Mayan communities [23], but not systematically mapped in terms of exposure.

Painted storefronts may provide for larger levels of exposure due to both the size of the advertisement relative to other marketing tactics, like posters and signs, and due to their permanence, given that stores stay painted with company logos and colors for years when compared to more rapidly changing bus stop and billboard ads and moving vehicles. Branded storefronts provide a large point-of-sale promotion since the stores bearing the brand’s colors and logos sell the company’s products on-site. The retailers also sell competitor sodas and other beverages, so painted storefronts may act as a priming device to nudge customers to choose the visible brand both at the time of buying drinks and during later purchasing decision-making.

The sort of visibility that painted storefronts afford in prime urban and rural locations would be worth thousands of dollars per month to building owners in countries like the United States. Some Guatemalan corner store owners have reported that the soda company whose brand their storefronts carried provided only the materials and labor involved in painting the buildings without additionally paying for ad space (personal communication with three store owners). More research is needed to confirm how widespread this lack of payment is, but the strategy does align with predatory practices that the UPF and SSB industry has been reported to employ *vis-a-vis* corner store owners in Guatemala [46].

### Home-based advertisements

Another previously unreported feature of the food environment we identify in Xela is ads for home-based sale of foods and beverages, which made up a fifth of all observed advertisements. Most modalities of commercial-grade advertisements raise brand awareness that, if acted on, may lead people to visit specific places to purchase foods and drinks, like fast-food restaurants, or select a specific brand when, at a later time, choosing from a selection of alternatives, such as in grocery stores and corner stores (*tiendas)*. Home-based ads are more localized and serve the purpose of indicating specific items for sale in a particular location. They have lower potential to reach visitors to the city and other passersby than commercial-grade ads because they are typically placed in less busy thoroughfares and are smaller in size than commercial-grade ads.

However, these types of less formal marketing are important to study in other locations from a public health policy and practice standpoint because they may supply city residents in specific neighborhoods, reaching the hyper-local community in which they are located (and possibly further afield by word of mouth). Home-based sale and advertising of foods, most of which people produce or prepare themselves, also underscores the continued importance of individual exchange of foodstuffs in many contexts. In Guatemala, a third of the population is employed in agriculture, while a much bigger proportion of the population, especially indigenous women, produces food for personal consumption and for sale from land they cultivate, animals they rear, and backyard patio production systems they maintain. This study has served as a starting point for estimating the contribution of home-based food and beverage advertising to the healthiness of retail food environments where non-corporate food provision from homes, eateries, and wet markets remains the norm, not an aberration [47].

### Public Health Implications

Through political activities, businesses in Guatemala have been documented to resist specific health-promoting food and beverage marketing policy reforms [48], which may contribute to the country’s very low ranking on nutrition policy implementation [49]. This is partly due to the revolving door between business leaders and political offices and the power the private sector wields in Guatemalan politics [50], which makes pro-public health regulation of commerce against the financial interests of the individuals and companies involved. This includes the country’s largest food and beverages companies and brands controlled by a handful of the wealthiest oligarchic families, including Pepsi bottling facilities, Embotelladora La Mariposa, and the national beer company, Gallo, both owned by the Castillo family; and the vertically-integrated fresh and fast chicken brands of Pollo Campero, Pinulito, and Pollo Rey that belong to Guatemala’s largest (family-owned) conglomerate, Corporación Multi Inversiones (CMI) owned by the Bosch-Gutiérrez family. However, the policy route to advancing public health nutrition in Guatemala is not completely closed as various domestic and transnational research groups have built the evidence base for introducing bills on labeling and food marketing [48]. As some note: “[f]or governments to act on unhealthy food advertising in outdoor spaces or on publicly owned assets, they must be equipped with the latest international evidence to support the need for policy and the technical design of regulation” [51]. This study contributes to such evidence-base building by measuring the total exposure to outdoor food and beverage ads in a busy area of an indigenous-majority city and evaluating the healthiness of the items being advertised.

Our Xela findings mark a starting point for building the evidence base for types and levels of exposure to outdoor food and beverage advertising. Future research in Guatemala could examine effects of outdoor advertising on diet preferences, food purchasing behavior, and health outcomes. It could also analyze policy options for reducing exposure to and any harmful effects of outdoor food and beverage advertising, and addressing predatory relationships between store owners and food and beverage industries.

This study also points the way to potential health-promoting actions. Practitioners could, for example, consider working with store owners to paint alternative storefront murals with messaging and visuals that promote public health nutrition. Public health marketers could consider counter-balancing the least healthy outdoor advertising messages with regular and visible promotions of more nutritious foods and drinks and the local retailers who provide them. Educators may also choose to educate the Guatemalan public about their outdoor food advertising environment to raise critical awareness of the kinds of messages they and their children are being exposed to.

Finally, these findings help counteract the negative imbalance of CDoH scholarship and the public health nutrition policy and practice implications arising from it. Despite calls to recognize both harms and benefits of commercial activities, existing research skews heavily towards negative CDoH by describing commerce’s “adverse health effects” and commercial activities as “drivers of ill-health” or “drivers of non-communicable diseases” [3, 6, 43]. The finding that none of the home-based ads for products that people grew or prepared themselves were rated as least healthy adds to the literature highlighting unregulated food and beverage enterprises as nutritional bright spots that contribute healthy options to retail food environments [52, 53].

### Strengths and Limitations

Contemporary food environments promote consumption of highly processed and energy-dense foods and drinks, encouraging deviation from traditional cuisines, particularly in less wealthy countries [54]. Studying outdoor food and beverage advertising as a CDoH in a demographic, educational, labor, and communications context like Xela’s sheds light on important drivers of and potential solutions to nutritional and epidemiological transitions around the world, especially among indigenous people. While one line of research has documented food and beverage marketing aimed at Guatemalan children [45, 55], one of this study’s strength lies in being the first to report on outdoor advertising of foods and beverages in an area affecting the general population. While most previous outdoor advertising research has considered a single modality, a single type of food or beverage, or a target location, such as schools, this paper estimated exposure from all food and beverage advertising across existing modalities in a given geographical space.

The present study’s results and conclusions are limited by the relatively small sample size of home-based and commercial-grade ads on the roads and streets that were mapped by a single researcher across multiple days before the COVID-19 pandemic. Further studies could systematically map all the streets in a given geographical area, expand the mapped area and compare across areas in order to increase the sample size, and conduct the mapping in a single day with multiple researchers to reduce possibility of measurement errors. Additional studies could determine whether the post-pandemic context has seen an increase or decrease in exposure to outdoor food and beverage advertisements of different healthiness in Xela.

Additionally, we did not survey all the residential streets and alleys in the mapped area. As a result, we likely under-sampled home-based advertising of foods and beverages, which were found more frequently on smaller residential streets and cul-de-sacs compared to commercial-grade ads that were concentrated on main thoroughfares. More research is needed to better estimate home-based ads’ contribution to the exposure to most healthy, middle healthy, and least healthy advertising people face in a given area in Guatemala and in other countries.

Our study did not take into account multiple in-store point of sale promotions that are visible from the street when passing by as some protocols do [56]. If it had, this kind of promotion would have likely counted upward of 450 additional food and beverage advertisements, most for unhealthy items or brands, fourfold outnumbering the 113 ads reported by this study. Future studies would need to consider whether or not to include point-of-sale promotional materials in corner stores that are visible from the street as part of outdoor food and beverage advertising.

Finally, since our study did not collect vehicle identifiers (like license plates), it is possible that some vehicles ads were double counted in our sample if the vehicles were observed more than once on different roads throughout the mapping period. While this double counting would inflate the number of unique vehicle-based ads in the study, it would not inflate the reported level of exposure to such ads from the viewpoint of a pedestrian on the street, which is what this study sought to measure. Future research would need to consider whether or not to include vehicle identifiers, depending on what the studies seek to monitor.

## Conclusions

Xela offers insights into some of the commercial activities contributing to the healthiness of retail food environments. This study’s findings widened the CDoH frame to consider unregulated food retail by individuals from their homes. The results also highlighted previously unreported business tactics to reach consumers through vehicle-based promotions and storefront branding in a context of deep employment, literacy, digital, and health inequities faced by indigenous people, especially women. The article has demonstrated that outdoor advertising is an important CDoH of central Xela’s retail food environment, with promotion of many least healthy items (a negative commercial determinant) but also promotion of some most healthy items (a positive commercial determinant). Future research and public health action might examine outdoor advertising’s links to diets and health outcomes, evaluate policy options for addressing predatory retail strategies, and consider public health initiatives that support nutritional bright spots.

## Data Availability

All relevant data are within the manuscript and its Supporting Information files. The dataset provided omits geolocation information for 21 home-based ads to protect individual residences from being identified.

## Acknowledgements

We thank the numerous in-country collaborators who were integral to the larger project that gave rise to this manuscript, including but not limited to representatives of organizations belonging to the El Colectivo Regional de Occidente (CORO), like Servicios Jurídicos y Sociales (SERJUS), Las Hojitas, Deutsche Gesellschaft für Internationale Zusammenarbeit (GIZ,) and Ministerio de Agricultura, Ganadería y Alimientación (MAGA). Paper writeup was made possible by the CDC’s inaugural Health Equity Science Manuscript Development Fellowship (henceforth, the Fellowship). We extend eternal gratitude to the fellows, UCSF faculty, and CDC staff involved in the Fellowship, who asked critical questions, made useful suggestions, and provided technical assistance during the conceptualization phase of the article. We are also grateful to CDC’s Prevention Research and Translation Branch (PRTB), especially to Brigette Ulin, Garry Lowry, and the rest of the Evaluation Team, for supporting the lead author with time release to participate in the Fellowship. Finally, this work benefited from critical reviews of related conceptual documents and early manuscript drafts by CDC’s Evelyn Twentyman, Stephanie Miedema, and Stephen Onufrak.

## APPENDIX A: Expert Rated Healthiness of Food and Beverage Items in the Functional Categories

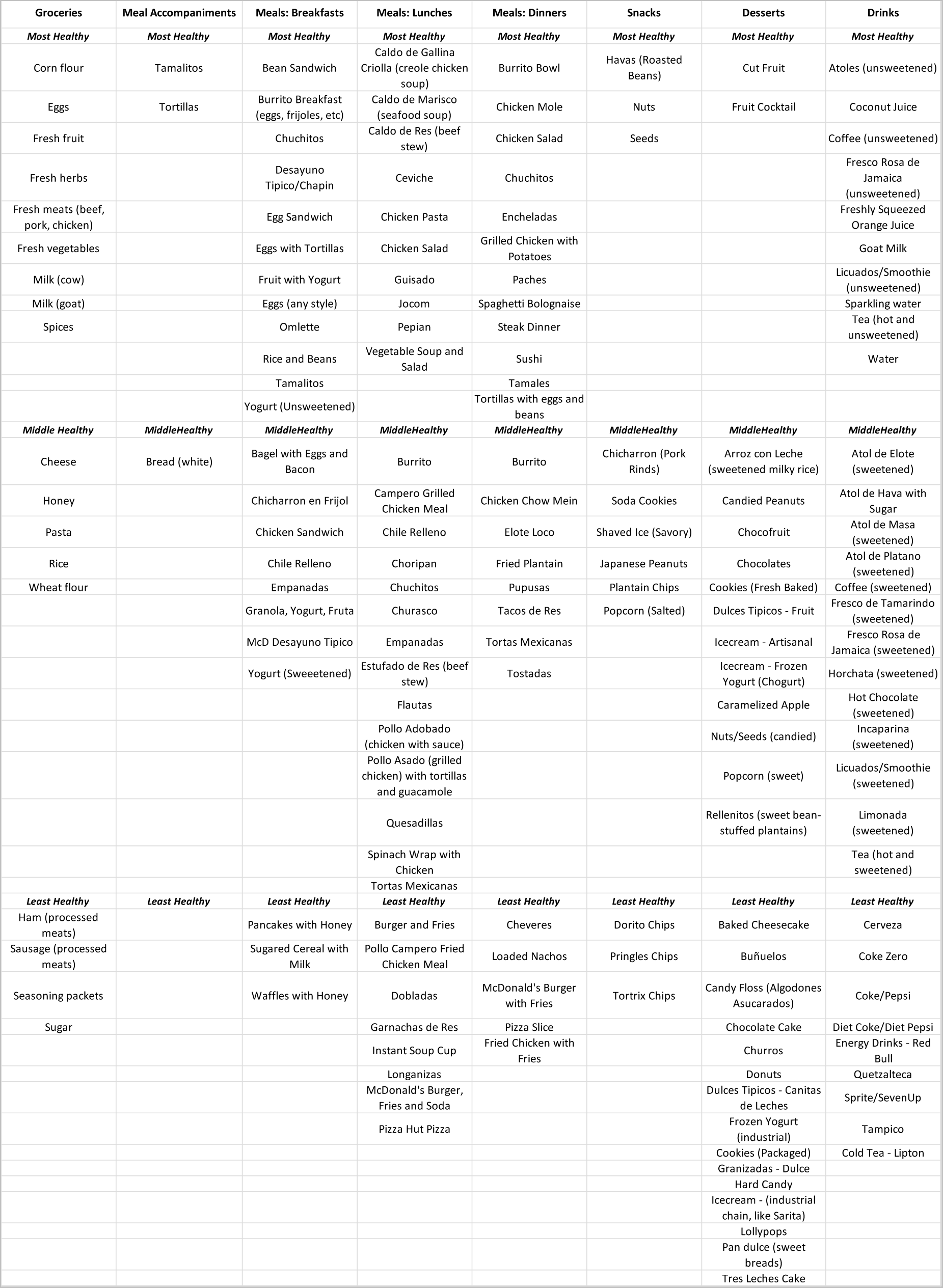

## Supporting Information

Supporting File 1: Data

